# Analysis of more than 400,000 women provides case-control evidence for BRCA1 and BRCA2 variant classification

**DOI:** 10.1101/2024.09.04.24313051

**Authors:** Maria Zanti, Denise G. O’Mahony, Michael T. Parsons, Leila Dorling, Joe Dennis, Nicholas J. Boddicker, Wenan Chen, Chunling Hu, Marc Naven, Kristia Yiangou, Thomas U. Ahearn, Christine B. Ambrosone, Irene L. Andrulis, Antonis C. Antoniou, Paul L. Auer, Caroline Baynes, Clara Bodelon, Natalia V. Bogdanova, Stig E. Bojesen, Manjeet K. Bolla, Kristen D. Brantley, Nicola J. Camp, Archie Campbell, Jose E. Castelao, Melissa H. Cessna, Jenny Chang-Claude, Fei Chen, Georgia Chenevix-Trench, NBCS Collaborators, Don M. Conroy, Kamila Czene, Arcangela De Nicolo, Susan M. Domchek, Thilo Dörk, Alison M. Dunning, A. Heather Eliassen, D. Gareth Evans, Peter A. Fasching, Jonine D. Figueroa, Henrik Flyger, Manuela Gago-Dominguez, Montserrat García-Closas, Gord Glendon, Anna González-Neira, Felix Grassmann, Andreas Hadjisavvas, Christopher A. Haiman, Ute Hamann, Steven N. Hart, Mikael B.A. Hartman, Weang-Kee Ho, James M. Hodge, Reiner Hoppe, Sacha J. Howell, kConFab Investigators, Anna Jakubowska, Elza K. Khusnutdinova, Yon-Dschun Ko, Peter Kraft, Vessela N. Kristensen, James V. Lacey, Jingmei Li, Geok Hoon Lim, Sara Lindström, Artitaya Lophatananon, Craig Luccarini, Arto Mannermaa, Maria Elena Martinez, Dimitrios Mavroudis, Roger L. Milne, Kenneth Muir, Katherine L. Nathanson, Rocio Nuñez-Torres, Nadia Obi, Janet E. Olson, Julie R. Palmer, Mihalis I. Panayiotidis, Alpa V. Patel, Paul D.P. Pharoah, Eric C. Polley, Muhammad U. Rashid, Kathryn J. Ruddy, Emmanouil Saloustros, Elinor J. Sawyer, Marjanka K. Schmidt, Melissa C. Southey, Veronique Kiak-Mien Tan, Soo Hwang Teo, Lauren R. Teras, Diana Torres, Amy Trentham-Dietz, Thérèse Truong, Celine M. Vachon, Qin Wang, Jeffrey N. Weitzel, Siddhartha Yadav, Song Yao, Gary R. Zirpoli, Melissa S. Cline, Peter Devilee, Sean V. Tavtigian, David E. Goldgar, Fergus J. Couch, Douglas F. Easton, Amanda B. Spurdle, Kyriaki Michailidou

## Abstract

Clinical genetic testing identifies variants causal for hereditary cancer, information that is used for risk assessment and clinical management. Unfortunately, some variants identified are of uncertain clinical significance (VUS), complicating patient management. Case-control data is one evidence type used to classify VUS, and previous findings indicate that case-control likelihood ratios (LRs) outperform odds ratios for variant classification. As an initiative of the Evidence-based Network for the Interpretation of Germline Mutant Alleles (ENIGMA) Analytical Working Group we analyzed germline sequencing data of *BRCA1* and *BRCA2* from 96,691 female breast cancer cases and 303,925 unaffected controls from three studies: the BRIDGES study of the Breast Cancer Association Consortium, the Cancer Risk Estimates Related to Susceptibility consortium, and the UK Biobank. We observed 11,227 *BRCA1* and *BRCA2* variants, with 6,921 being coding, covering 23.4% of *BRCA1* and *BRCA2* VUS in ClinVar and 19.2% of ClinVar curated (likely) benign or pathogenic variants. Case-control LR evidence was highly consistent with ClinVar assertions for (likely) benign or pathogenic variants; exhibiting 99.1% sensitivity and 95.4% specificity for *BRCA1* and 92.2% sensitivity and 86.6% specificity for *BRCA2*. This approach provides case-control evidence for 785 unclassified variants, that can serve as a valuable element for clinical classification.

## Introduction

Clinical genetic testing of disease-associated susceptibility genes is often complicated by the identification of variants of uncertain clinical significance (VUS)^1^. These individually rare variants may include missense changes, intronic and small in-frame insertions or deletions, as well as regulatory variants for which the association with disease is uncertain, complicating counselling and clinical management^2^.

To facilitate the classification of variants identified by genetic testing into (likely) benign or (likely) pathogenic, the American College of Medical Genetics and Genomics and the Association for Molecular Pathology (ACMG/AMP) groups^3^ developed guidelines that incorporate a set of criteria representing different evidence types. In this framework, independent lines of evidence in favor of or against pathogenicity for a variant are weighted according to strength level, where possible based on likelihood ratios (LRs), and following recommendations based on Bayesian modeling of the ACMG/AMP criteria^4^. Specific guidelines for *BRCA1* and *BRCA2*, based on this framework, have been published by the ClinGen Evidence-based Network for the Interpretation of Germline Mutant Alleles (ENIGMA) *BRCA1* and *BRCA2* Variant Curation Expert Panel (VCEP)^2^. Classification of a variant as (likely) benign or (likely) pathogenic (or remaining as VUS) is determined by joint assessment of the evidence types, either according to the combination of criteria met^3,4^ or following a points-scoring system recently developed^5^. Based on previous research by the ENIGMA consortium^6^, one evidence type adapted for use under the ACMG/AMP framework is that from case-control data. In the ACMG/AMP framework this was included in the PS4 criterion, defined as “the prevalence of the variant in affected individuals is significantly increased over controls, with advice on application that the relative risk (RR) or odds ratio (OR) from case-control studies should be > 5.0, and the confidence interval (CI) around the estimate should not include 1.0”^3^. ClinGen specifications of the ACMG/AMP framework for *BRCA1* and *BRCA2* suggest that an OR ≥ 4.0 is assigned at full strength for a statistically significant association with CI not including 2.0^2^. As an initiative of the ENIGMA Analytical Working Group, we have recently proposed a LR-based framework for the analysis of case-control data for variant classification, where derived LRs are applicable under the ACMG/AMP framework for variant classification^7^. Compared to ORs derived by logistic regression analysis, the LR-based framework has vastly improved performance to provide evidence towards pathogenicity, and more importantly, it can also be used to derive evidence against pathogenicity. For this reason, the BRCA1/2 VCEP specifications also state that ccLR estimates should be used in in preference to case-control OR data for application of case-control information (https://cspec.genome.network/cspec/ui/svi/).

Herein, comparing the case-control LR (ccLR) method and logistic regression analysis, we performed a large-scale assessment of 4,302 unique variants in *BRCA1* (2,254 within coding sequence (CDS) ±5bp) and 6,925 unique variants in *BRCA2* (4,667 CDS±5bp) based on sequencing data from 96,691 female breast cancer cases and 303,925 unaffected female controls from three cohorts: the BRIDGES project of the Breast Cancer Association Consortium (BCAC)^8^, the Cancer Risk Estimates Related to Susceptibility (CARRIERS) consortium^9^ and the UK Biobank (UKB)^10^.

## Results

Case-control LRs were aligned to evidence strength levels for or against pathogenicity based on the thresholds recommended under the ACMG/AMG framework^4^. To evaluate the effect of variant counts on the reliability of the results, we performed analyses considering variants that were present in at least one, two or three individuals in each dataset separately (**Supplementary Tables 1-2**) and in the combined dataset (**Supplementary Table 3**). To maintain low false discovery and false omission rates (< 0.05) and reach strong evidence in favor and against pathogenicity for ccLR estimate application (**Supplementary Tables 1-3**), results indicated that a more conservative approach would require variants present in at least three individuals in the combined dataset (*BRCA1* and *BRCA2*) and evidence from at least two datasets (only for *BRCA2*). By applying these specifications, the enrichment ratios for known pathogenic:benign variants in each ccLR evidence category were broadly consistent with the expectation for that category, with the exception of *BRCA2* where ccLR benign supporting evidence fell into the “No evidence” category (**Supplementary Table 3**, **Table 1**).

**Table 1.** Likelihood ratios (LRs) towards pathogenicity for (likely) benign and (likely) pathogenic variants assigned ccLR ACMG/AMP evidence strengths.

|  | (Likely)<br>Benign |  | (Likely)<br>Pathogenic |  | LR towards<br>pathogenicity | LCI | HCI | ACMG/AMP<br>Evidence strength |
| --- | --- | --- | --- | --- | --- | --- | --- | --- |
|  | N | Prop | N | Prop |  |  |  |  |
| **BRCA1 reference set** |  |  |  |  |  |  |  |  |
| Pathogenic Strong | 0 | 0.000 | 55 | 0.585 | 288.600 | 18.008 | 4625.248 | Pathogenic Strong |
| Pathogenic Moderate | 2 | 0.008 | 21 | 0.223 | 27.479 | 6.571 | 114.920 | Pathogenic Strong |
| Pathogenic Supporting | 2 | 0.008 | 7 | 0.074 | 9.160 | 1.937 | 43.303 | Pathogenic Moderate |
| No evidence | 27 | 0.110 | 9 | 0.096 | 0.872 | 0.426 | 1.785 | No evidence |
| Benign Supporting | 23 | 0.093 | 0 | 0.000 | 0.055 | 0.003 | 0.902 | Benign Moderate |
| Benign Moderate | 43 | 0.175 | 1 | 0.011 | 0.061 | 0.009 | 0.436 | Benign Moderate |
| Benign Strong | 149 | 0.606 | 1 | 0.011 | 0.018 | 0.002 | 0.124 | Benign Strong |
| **Total variants identified in dataset** | **246** |  | **94** |  |  |  |  |  |
| **BRCA2 reference set** |  |  |  |  |  |  |  |  |
| Pathogenic Strong | 1 | 0.004 | 50 | 0.397 | 110.317 | 15.411 | 789.691 | Pathogenic Strong |
| Pathogenic Moderate | 8 | 0.029 | 20 | 0.159 | 5.516 | 2.497 | 12.183 | Pathogenic Moderate |
| Pathogenic Supporting | 4 | 0.014 | 14 | 0.111 | 7.722 | 2.593 | 22.993 | Pathogenic Moderate |
| No evidence | 34 | 0.122 | 22 | 0.175 | 1.428 | 0.872 | 2.338 | No evidence |
| Benign Supporting | 22 | 0.079 | 8 | 0.063 | 0.802 | 0.367 | 1.753 | No evidence |
| Benign Moderate | 47 | 0.169 | 9 | 0.071 | 0.422 | 0.214 | 0.835 | Benign Supporting |
| Benign Strong | 162 | 0.583 | 3 | 0.024 | 0.041 | 0.013 | 0.126 | Benign Strong |
| **Total variants identified in dataset** | **278** |  | **126** |  |  |  |  |  |

Reference sets included (likely) pathogenic and (likely) benign variants located within coding sequence (CDS) or splice acceptor/donor site dinucleotide positions ( $\pm 2$ bp), classified as such by the ClinGen *BRCA1/2* historical expert panel or the ClinGen *BRCA1/2* VCEP following ACMG/AMP guidelines, or by multiple submitters without conflicts in the ClinVar database, and also with gnomAD (v4.1.0) maximum credible allele frequency in the non-founder populations $\leq 0.001$ consistent with the *BRCA1* and *BRCA2* VCEP's proposed BA1 cutoff. For likelihood ratio estimation, the Haldane-Anscombe correction was applied for any category where the cell count for pathogenic or benign variant reference set was zero. *Code weights based on LRs with confidence intervals spanning 1 are shown in italics.*

The combined dataset of 96,691 female breast cancer cases and 303,925 unaffected female controls from BRIDGES, CARRIERS and the UKB contained case-control counts for 11,227 variants. Specifically, 4,302 variants were found in *BRCA1* and 6,925 in *BRCA2.* The dataset encompasses 21.3% (6,510/30,609) of the ClinVar *BRCA1* and *BRCA2* curated variants, 19.2% (2,991/15,565) of the ClinVar variants curated as (likely) benign or pathogenic, and 23.4% (3,519/15,044) of the ClinVar variants annotated as VUS.

When focusing solely on exonic and proximal intronic sequences (CDS±5bp); covered adequately by all three cohorts, the dataset represented 18.7% (5,729/30,609) of the ClinVar database and 21.9% (3,301/15,044) of the ClinVar VUS. Of the total CDS±5bp variants, there are 2,254 variants in *BRCA1* and 4,667 in *BRCA2*. Further filtering for maximum credible AF in the non-founder populations (gnomAD v4.1.0), denoted as filtering allele frequency (FAF) ≤ 0.001 (*BRCA1* and *BRCA2* VCEP “BA1” benign stand-alone cutoff), present in at least three individuals in the combined dataset, and with evidence from at least two datasets for *BRCA2*, led to a final set of 1,717 variants (685 in *BRCA1* and 1,032 in *BRCA2*). These included: 765 variants listed in ClinVar as (likely) pathogenic or (likely) benign; and 952 variants considered to be of uncertain significance because they were not reported in ClinVar or listed in ClinVar as VUS or with conflicting interpretation of pathogenicity (**Fig. 1a**). Sequence ontology variant consequence was annotated as follows: 207 presumed loss-of-function (LOF) variants of which 129 were frameshift insertions or deletions, 23 canonical splice site (±2bp), 55 nonsense variants, as well as 31 in-frame insertions or deletions, 369 synonymous, 1,084 missense and 26 intronic (±3 to ±5bp) variants (**Fig. 1b**). Case-control LRs provided evidence in favor of pathogenicity for 266 variants and against pathogenicity for 1,187 variants; of these 1,453 variants, 785 were considered to be of uncertain significance (**Fig. 1c**). Case-control results for the final set of 1,717 *BRCA1* and *BRCA2* variants, are summarized and displayed as genomic maps in **Figure 2 and Figure 3**.

**Fig. 1:**
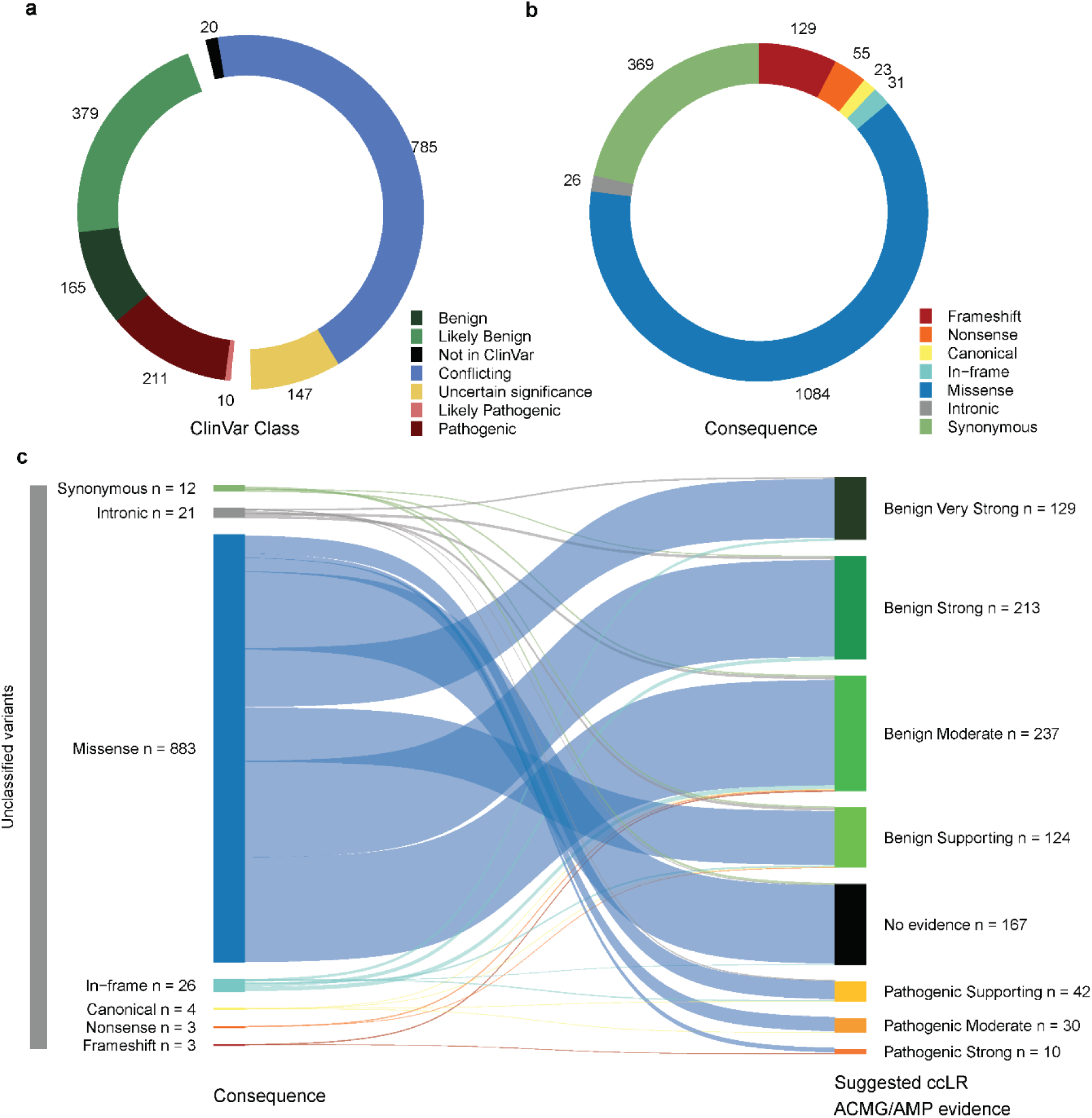
Overview of variants with provided case-control likelihood ratio evidence. Donut plots showing the distribution of the **a** clinical classification (“ClinVar Class”) and **b** sequence ontology variant consequence (“Consequence”) for the 1,717 CDS±5bp variants with filtering allele frequency (FAF) > 0.001 (variants not meeting the *BRCA1* and *BRCA2* VCEP “BA1” benign stand-alone criterion), present in at least three individuals in the combined dataset, and with evidence from at least two datasets for *BRCA2*. **c** Sankey plot depicting “suggested case-control likelihood ratio (ccLR) ACMG/AMP evidence” provided for unclassified variants (not reported in ClinVar or listed in ClinVar as VUS, variants of conflicting interpretation of pathogenicity or variants with classification “not provided”), per sequence ontology variant consequence (“Consequence”). The clinical classification status (“ClinVar Class”) of variants was retrieved from the ClinVar database (last accessed on January 7, 2024).

**Fig. 2:**
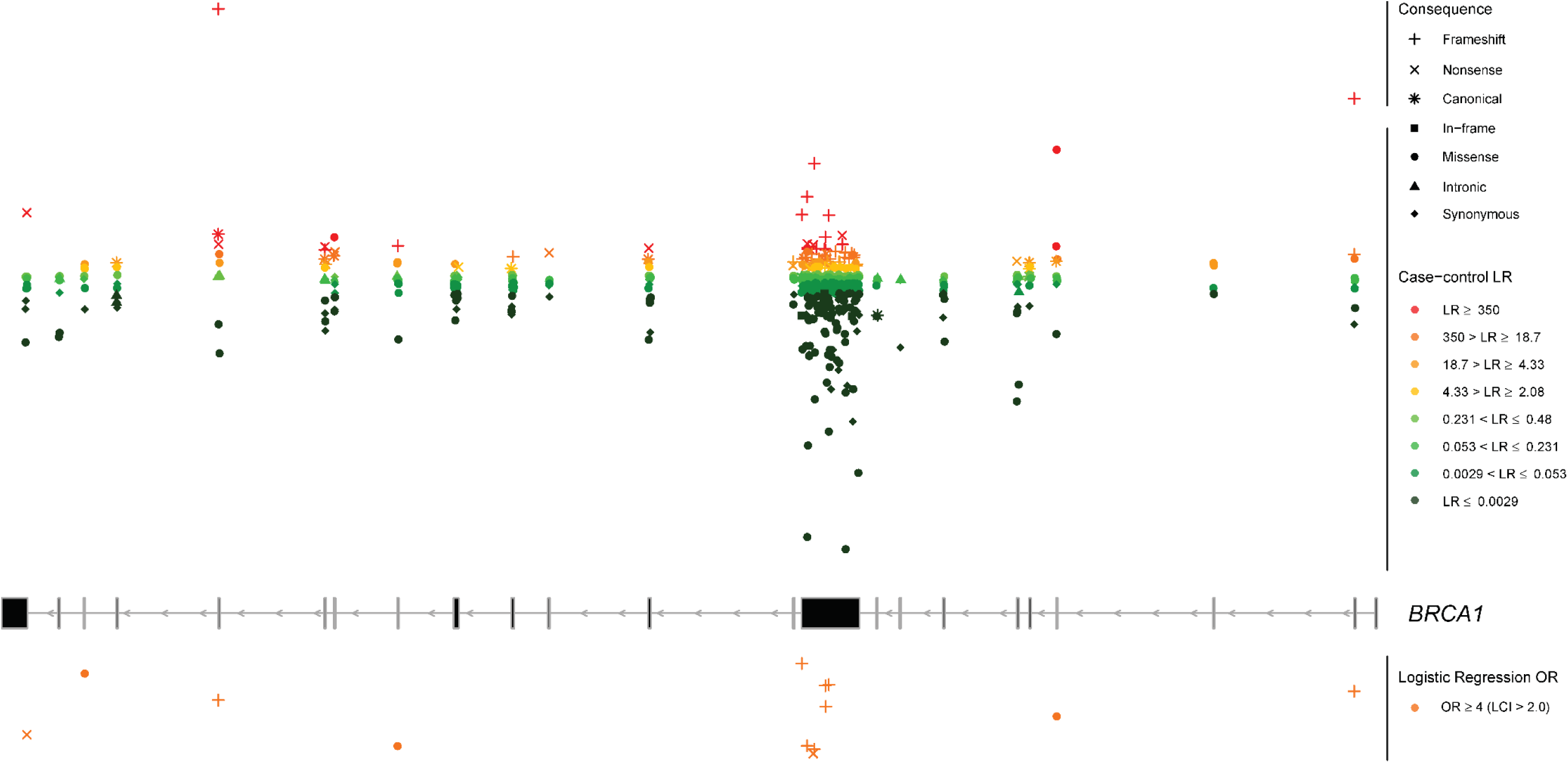
Genomic mapping of the case-control analysis for *BRCA1*. Overlay of the case-control likelihood ratios (LRs) and the logistic regression odds ratio (OR) estimates is represented within each exon (middle panel). Exons are sequentially numbered from 1 to 23 and annotated from right to left to match the MANE Select transcripts. Although *BRCA1* was initially described with 24 exons (GenBank Assession ID U14680.1), exon 4 is missing following further assessment of the gene. We implement the most updated version of exon numbering (excluding legacy exon numbering). Case-control LRs (top panel) are represented on a continuous log2-transformed y axis with axis breaks. For the case-control LR analysis, red color gradient represents LR reaching suggested ACMG/AMP evidence in favor of pathogenicity with strength levels ranging from very strong (dark red), to supporting (yellow). Green color gradient represents LR reaching ACMG/AMP evidence against pathogenicity with strength levels ranging from very strong (dark green) to supporting (light green). Variants with LR of “No evidence” are not plotted. For the logistic regression analysis, orange color represents OR estimates reaching the strong PS4 criterion (OR ≥ 4.0, P value < 0.05 and confidence interval (CI) not including 2.0). Variants with OR estimates not reaching the PS4 criterion are not plotted. For visualization purposes, the y axis for logistic regression is represented in reversed order. LCI, lower confidence interval. Sequence ontology variant consequence (“Consequence”) is represented with different symbols.

**Fig. 3:**
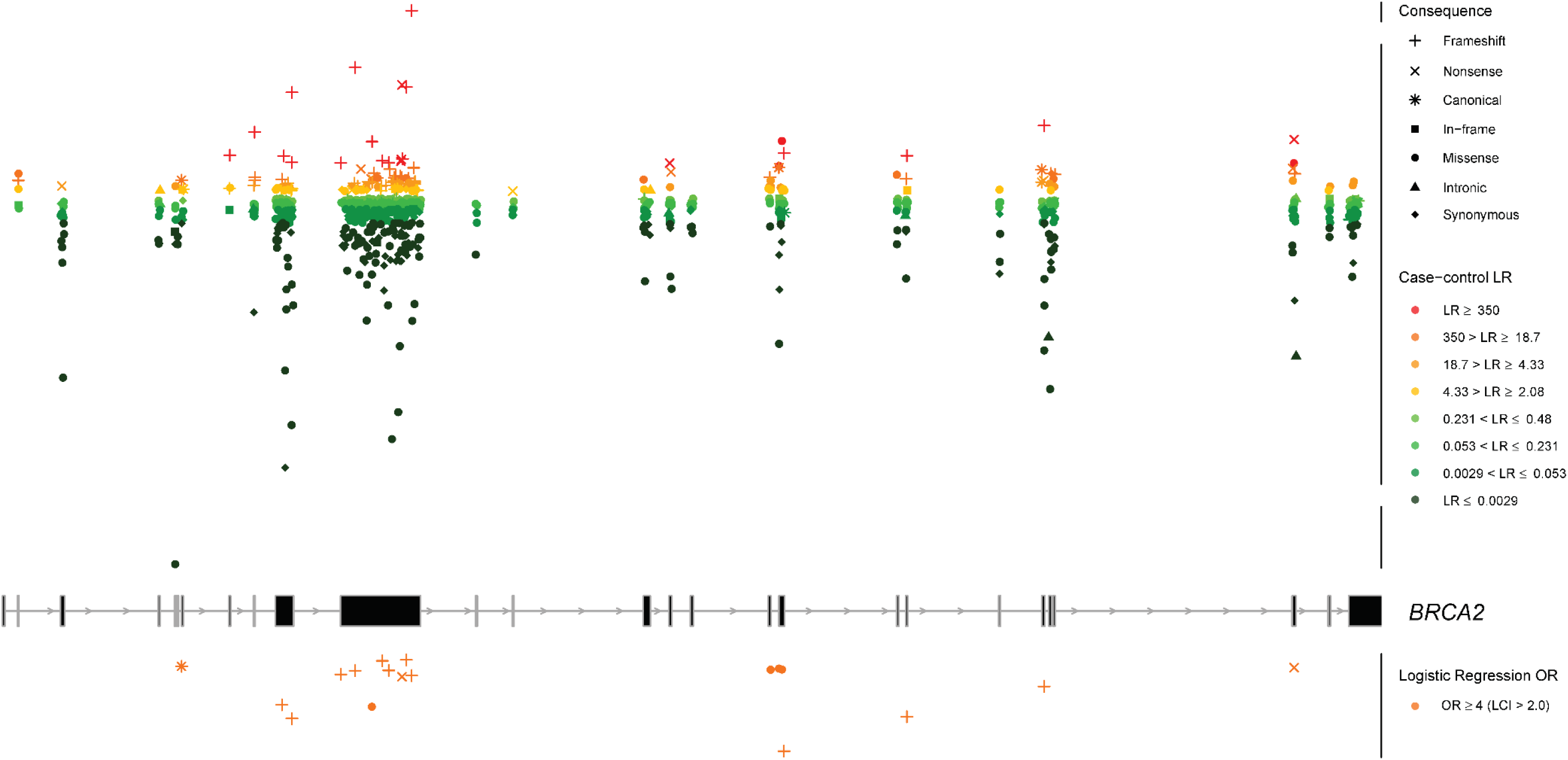
Genomic mapping of the case-control analysis for *BRCA2*. Overlay of the case-control likelihood ratios (LRs) and the logistic regression odds ratio (OR) estimates is represented within each exon (middle panel). Exons are sequentially numbered from 1 to 27 and annotated from left to right to match the MANE Select transcripts. Case-control LRs (top panel) are represented on a continuous log2-transformed y axis with axis breaks. For the case-control LR analysis, red color gradient represents LR reaching suggested ACMG/AMP evidence in favor of pathogenicity with strength levels ranging from very strong (dark red), to supporting (yellow). Green color gradient represents LR reaching ACMG/AMP evidence against pathogenicity with strength levels ranging from very strong (dark green) to supporting (light green). Variants with LR of “No evidence” are not plotted. For the logistic regression analysis, orange color represents OR estimates reaching the strong PS4 criterion (OR ≥ 4.0, P value < 0.05 and confidence interval (CI) not including 2.0. Variants with OR estimates not reaching the PS4 criterion are not plotted. For visualization purposes, the y axis for logistic regression is represented in reversed order. LCI, lower confidence interval. Sequence ontology variant consequence (“Consequence”) is represented with different symbols.

### BRCA1 Likelihood Ratios

Informative case-control LRs were obtained for 604 out of 685 CDS±5bp *BRCA1* variants (88.2%) (**Fig. 4**). Among these, evidence in favor of pathogenicity was observed for 116 variants; of those 27 were categorized as very strong, 30 as strong, 34 as moderate and 25 as supporting. Conversely, evidence against pathogenicity was provided for 488 variants; 156 categorized as very strong, 129 as strong, 119 as moderate, and 84 as supporting. In comparison, using logistic regression analysis only 13 variants reached strong evidence in favor of pathogenicity according to the *BRCA1/2* ACMG/AMP classification criterion description for OR application (PS4 criterion, OR ≥ 4.0, *P* value < 0.05 and CI not including 2.0)^2^, and all of these were also assigned pathogenic evidence using the ccLR method.

**Fig. 4:**
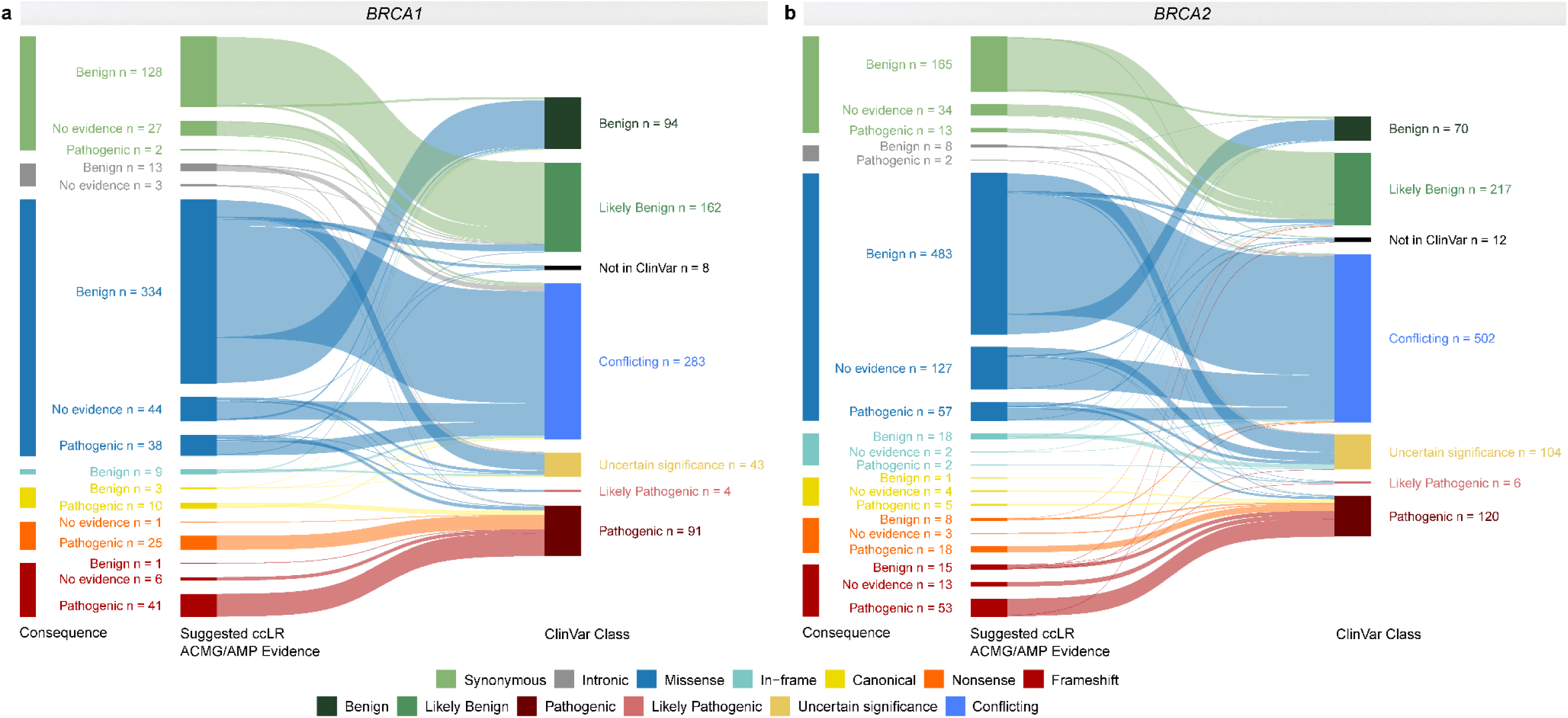
Case-control likelihood ratio evidence per sequence ontology variant consequence compared to ClinVar clinical classification. Sankey plots for a *BRCA1* and b *BRCA2*. The variants assigned case-control likelihood ratio (LR) evidence in favor of or against pathogenicity (with suggested supporting, moderate, strong or very strong evidence strength) are simplistically annotated as “Pathogenic”, and “Benign” “Suggested case-control LR (ccLR) ACMG/AMP Evidence”, respectively. Variants with LRs between 0.48 and 2.08 are defined as “No evidence” in the “Suggested ccLR ACMG/AMP Evidence” panel. The clinical classification status (“ClinVar Class”) of variants was retrieved from the ClinVar database (last accessed on January 7, 2024).

Variants having case-control LR evidence in favor of pathogenicity consisted of 65.5% (76/116) LOF (41 frameshift insertions or deletions, 10 canonical splice site and 25 nonsense variants). Missense and synonymous variants accounted for 32.8% (38/116) and 1.7% (2/116), respectively. In contrast, variants with evidence against pathogenicity included 0.8% (4/488) LOF (1 frameshift, and 3 canonical splice site variants), 1.8% (9/488) in-frame, 68.4% (334/488) missense, 2.7% (13/488) intronic and 26.2% (128/488) synonymous variants (**Fig. 4** and **Fig. 5a**). Among the 13 variants reaching the PS4 criterion, 76.9% (n = 10) were LOF (8 frameshift and 2 nonsense variants) alongside 23.1% (n = 3) missense variants. For *BRCA1* LOF variants, pathogenicity was provided for the majority of the frameshift insertions or deletions (97.6%, 41/42), canonical splice site (76.9%, 10/13) and nonsense (100%, 25/25) variants with informative case-control evidence.

**Fig. 5:**
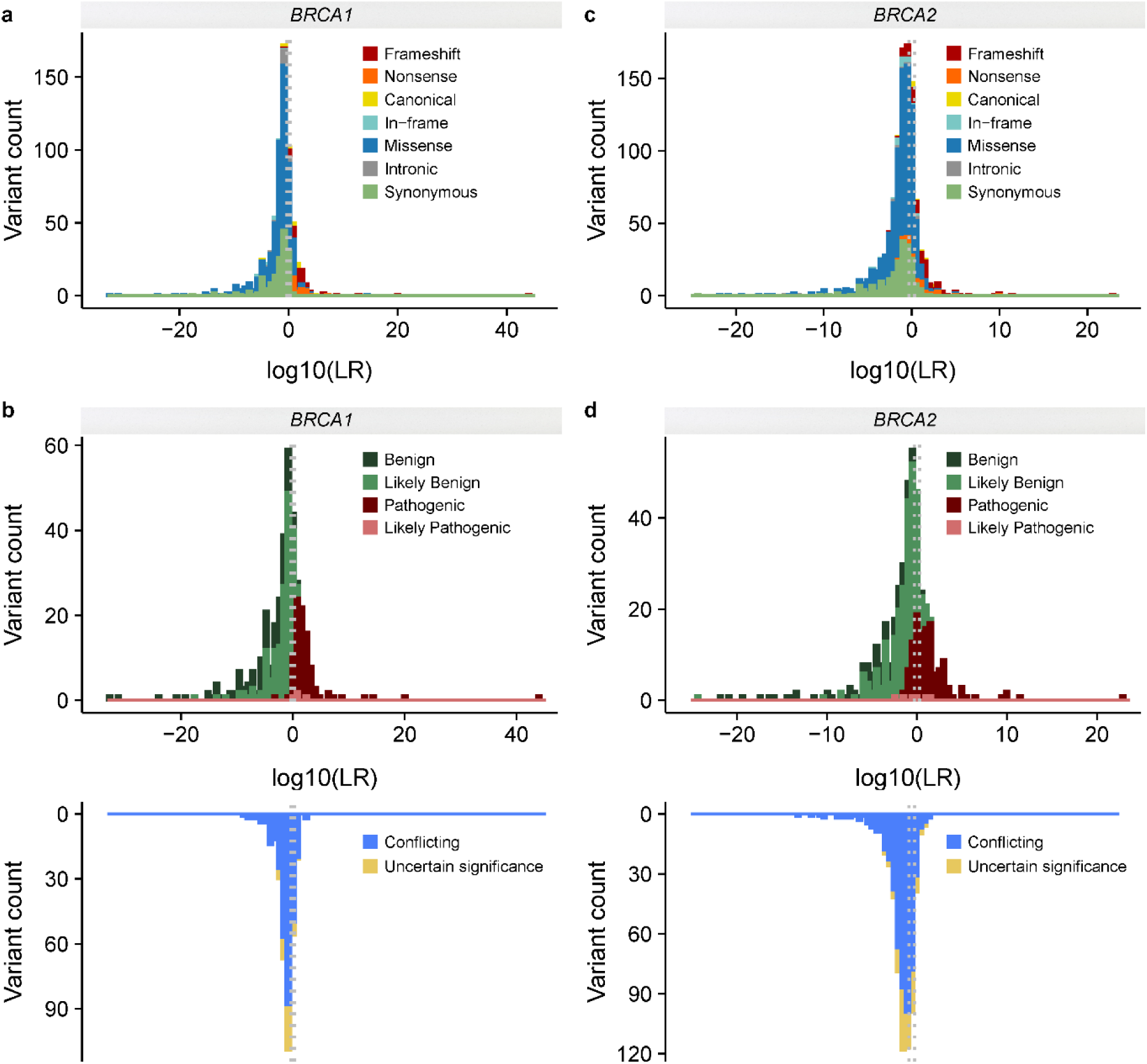
Distribution of the case-control likelihood ratios for *BRCA1* and *BRCA2*. Histograms showing the distribution of case-control likelihood ratios (LRs) categorized by **a** sequence ontology variant consequence for *BRCA1*, **b** ClinVar classification for *BRCA1*, **c** sequence ontology variant consequence for *BRCA2*, **d** ClinVar classification for *BRCA2*. For visualization purposes the x axis represents log10(LR) values. Dashed lines represent LRs between 0.48 and 2.08 considered as of “No evidence”.

The proportion of variants with informative case-control evidence towards pathogenicity was 10.2% (38/372) for missense, and 1.5% (2/130) for synonymous variants. All the *BRCA1* in-frame (n = 9) and intronic (n = 13) variants with informative case-control evidence were assigned benign evidence (**Fig. 4**). The majority of the identified CDS±5bp *BRCA1* variants were within exon 10 (n = 423), followed by exons 15 (n = 30) and 14 (n = 21). One splice acceptor (c.594-2A>C) and two splice donor (c.301+1G>A and c.4096+1G>A) variants within or proximal to exons 8, 5 and 10 were assigned benign evidence. Furthermore, missense variants with evidence in favor of pathogenicity were enriched in the exons encoding the RING domain (nucleotides 4-294; exons 2-5) (18.4%, 7/38) and the BRCA1-C-Terminal (BRCT) repeats (nucleotides 4987-5577, exons 16-23) (26.3%, 10/38) (**Fig. 6**). None of the in-frame variants assigned benign case-control (n = 9) evidence fell within the RING or BRCT domains.

**Fig. 6:**
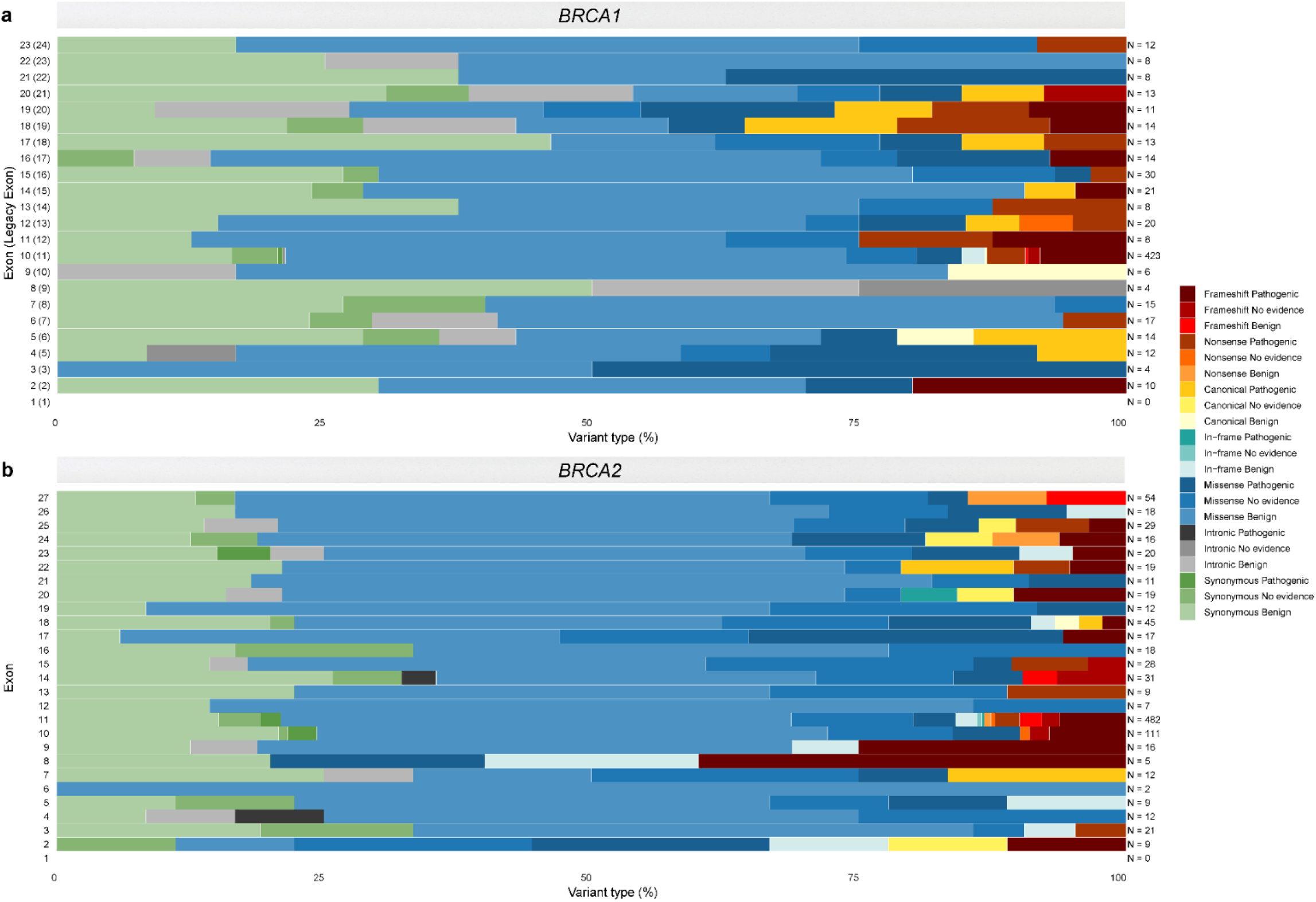
Overview of the case-control likelihood ratio evidence assigned per exon and sequence ontology variant consequence for *BRCA1* and *BRCA2*. Stacked bar plots of the suggested case-control likelihood ratio (LR) ACMG/AMP evidence per exon and sequence ontology variant consequence for **a** *BRCA1* and **b** *BRCA2.* Exons are sequentially numbered to match the MANE Select transcripts. Although *BRCA1* was initially described with 24 exons (GenBank Assession ID U14680.1), exon 4 is missing following further assessment of the gene; legacy exon numbering for *BRCA1* is represented in brackets. Variants assigned case-control LR evidence in favor of or against pathogenicity (with suggested supporting, moderate, strong or very strong evidence strength) are simplistically annotated as “Pathogenic”, and “Benign” in the key, respectively. Variants with LRs between 0.48 and 2.08 are defined as “No evidence”.

Overall, 25% (29/116) of the variants with evidence in favor of pathogenicity and 53.7% (262/488) of the variants with assigned evidence against pathogenicity, were considered to be of uncertain significance prior to evaluation (not reported in ClinVar, or listed in ClinVar as VUS, with conflicting interpretation of pathogenicity, or with classification “not provided”). For the remaining variants with an established ClinVar clinical classification as (likely) benign or (likely) pathogenic, we observed 99.1% (224/226) consistency for variants assigned benign evidence and 95.4% (83/87) for variants assigned pathogenic evidence (**Fig. 4** and **Fig. 5b**). For the 13 variants that reached the PS4 criterion using logistic regression analysis, one was of uncertain significance and the remainder had an established ClinVar clinical classification consistent with PS4 evidence in favor of pathogenicity.

### BRCA2 Likelihood Ratios

Informative case-control LRs were obtained for 82.3% of the *BRCA2* variants (849/1,032) (**Fig. 4**). Evidence in favor of pathogenicity was provided for 150 variants, with 24 being very strong, 35 strong, 47 moderate and 44 supporting. Evidence against pathogenicity was provided for 699 variants, of which 181 reached very strong, 195 strong, 225 moderate and 98 supporting. Using logistic regression analysis only 18 variants reached strong evidence in favor of pathogenicity following the *BRCA1/2* ACMG/AMP PS4 classification criterion (OR ≥ 4.0, *P* value < 0.05 and CI not including 2.0); all of these were assigned pathogenic evidence using the ccLR method.

Variants assigned ccLR evidence in favor of pathogenicity comprised of 50.7% (76/150) loss-of-function (53 frameshift insertions or deletions, 18 nonsense and 5 canonical splice site variants), 38% (57/150) missense, 8.7% (13/150) synonymous, 1.3% (2/150) intronic and 1.3% (2/150) in-frame variants. In contrast variants assigned ccLR evidence against pathogenicity comprised of 3.4% (24/699) loss-of-function (15 frameshift, 8 nonsense and 1 canonical splice site variants), 2.6% (18/699) in-frame, 69.2% (484/699) missense, 1.1% (8/699) intronic and 23.6% (165/699) synonymous variants. The 18 variants reaching the PS4 OR criterion were comprised of 14 loss-of-function (77.8%) (11 frameshift insertions or deletions, 2 nonsense and 1 canonical splice site variants) and 4 (22.2%) missense variants (**Fig. 4** and **Fig. 5a**). For *BRCA2* LOF variants, we observed that pathogenicity was proposed for the majority of the frameshift insertions or deletions (77.9%, 53/68), canonical splice site (83.3%, 5/6) and nonsense (69.2%, 18/26) variants assigned informative case-control evidence. The proposed pathogenicity rate for non-LOF variants (in-frame, missense, synonymous and intronic variants) with informative case-control evidence, was 10% (2/20) for in-frame, 10.5% (57/541) for missense, 7.3% (13/178) for synonymous and 20% (2/10) for intronic variants (**Fig. 4**). The majority of the evaluated *BRCA2* variants were within exons 11 (n = 482) and 10 (n = 111) (**Fig. 6**). Two intronic variants (c.425+3A>G, c.7435+5T>C) were assigned evidence in favor of pathogenicity, falling within the splice donor sites of exons 4 and 14. Moreover, two in-frame variants within exons 11 and 20 were assigned evidence in favor of pathogenicity. One splice donor variant (c.8331+2T>C) in the splice donor site of exon 18 was assigned evidence against pathogenicity. Finally, missense variants assigned evidence in favor of pathogenicity were enriched in the exons encoding the BRCA DNA binding domain (DBD) (amino acids 2481-3186; exons 15-26) (36.8%, 21/57) (**Fig. 6**).

Overall, 35.3% (53/150) of the variants with evidence in favor of pathogenicity and 63.1% (441/699) with assigned evidence against pathogenicity, were considered to be of uncertain significance prior to evaluation (not reported in ClinVar, or listed in ClinVar as VUS, with conflicting interpretation of pathogenicity, or with classification “not provided”). For the remaining variants with an established ClinVar clinical classification as (likely) benign or (likely) pathogenic, we observed 92.2% (238/258) consistency for variants assigned benign evidence and 86.6% (84/97) for variants assigned pathogenic evidence (**Fig. 4** and **Fig. 5b**). For the 18 variants that met the PS4 OR criterion, one was of uncertain significance and the remainder had PS4 evidence consistent with an established ClinVar clinical classification.

### Concordance with predicted or experimental impact on function

Concordance with evidence in favor or against pathogenicity using the ccLR method was higher for experimental evidence from higher-throughput functional assays compared to *in silico* prediction methods (**Supplementary Table 4**). We observed 72.4% sensitivity and 98.7% specificity for functionally evaluated variants located in 13 exons of *BRCA1* encoding the RING and BRCT domains (exons 2-5 and 15-23, respectively)^11^ and 100% sensitivity and 93.3% specificity for functionally evaluated missense *BRCA2* variants^12^. We also observed a strong concordance between the ccLR method and the recent functional evaluation of variants located in the region encoding the C-terminal DNA binding domain (DBD) of *BRCA2*^13^, with 64.3% sensitivity and 92.1% specificity. For *in silico* prediction methods evaluated, including BayesDel^14^, REVEL^15^, VEST4^16^, MutPred2^17^ and AlphaMissense^18^, we observed a moderate concordance with sensitivity ranging from 29.7% to 62.5%, and specificity ranging from 69% to 96.2% for *BRCA1*, and sensitivity ranging from 13.2% to 30.3% and specificity ranging from 88.2% to 95.1% for *BRCA2* (**Fig. 7** and **Supplementary Table 4**).

**Fig. 7:**
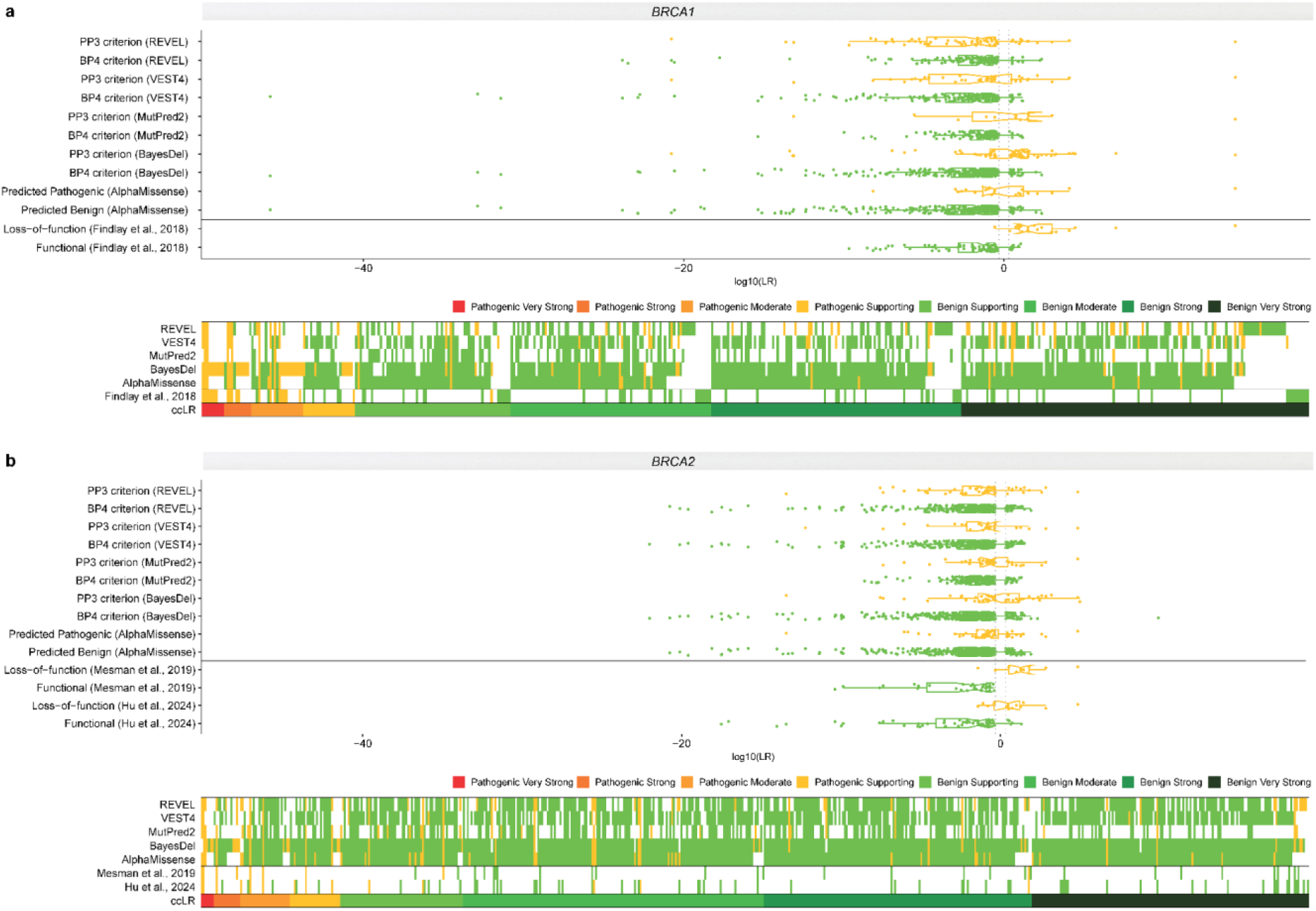
Concordance between the case-control likelihood ratio method and functional predictors. Concordance is shown separately for **a** *BRCA1* and **b** *BRCA2*. The top panels for each gene represent case-control likelihood ratios (LRs) compared to variants predicted as benign (“BP4 criterion”, “predicted benign” or “functional”) or pathogenic (“PP3 criterion”, “predicted pathogenic” or “loss-of-function”) by *in silico* prediction methods (AlphaMissense, BayesDel, MutPred2, VEST4 and REVEL) or through high-throughput functional assays (Findlay et al., 2018, Hu et al., 2024, Mesman et al., 2019). Yellow and green colors represent variants predicted as pathogenic or benign by functional predictors, respectively. Bottom panels for each gene represent sequence-pathogenicity heatmaps demonstrating the concordance between the case-control LR (ccLR) method and functional predictors. For the case-control LR evidence, red color gradient represents LR reaching suggested ACMG/AMP evidence in favor of pathogenicity with strength levels ranging from very strong (dark red), to supporting (yellow). Green color gradient represents LR reaching suggested ACMG/AMP evidence against pathogenicity with strength levels ranging from very strong (dark green) to supporting (light green). Variants with ccLR of “No evidence” are not plotted. For the functional predictors, yellow and green colors represent evidence in favor and against pathogenicity, respectively (expressed as pathogenic supporting).

Taken together, high-throughput assay results align with ccLR evidence, offering a robust evaluation framework. Concordance between the ccLR method and predicted or experimental impact is displayed as sequence-pathogenicity heatmaps (**Fig. 7**) which provide pathogenicity predictions for overlapping variants and functional evidence (binary categories) with ccLR.

## Discussion

Significant efforts have been dedicated to the clinical classification of variants in *BRCA1* and *BRCA2*, owing to their elevated risk association with multiple cancer types. To date, only 111 *BRCA1* and *BRCA2* variants have been assigned case-control LR evidence^7,19^, and only 20 of these were previously used in clinical classification^19^. In this work, we present a large-scale multicenter case-control analysis of 11,227 *BRCA1* and *BRCA2* rare variants of which 6,921 are coding (within CDS±5bp). Using sequencing data from 96,691 female breast cancer cases and 303,925 unaffected controls of the BRIDGES, CARRIERS and the UK Biobank datasets we utilized our recently developed ccLR method^7^ and logistic regression analysis to leverage case-control data and provide evidence in favor or against variant pathogenicity. Our dataset comprehensively covers all exons, proximal intronic sequences and regulatory regions, accounting for 21.3% of the ClinVar curated variants and 23.4% of the ClinVar VUS.

For coding variants, case-control evidence strongly corresponds with ClinVar pathogenicity data, exhibiting 99.1% sensitivity and 95.4% specificity in distinguishing (likely) pathogenic and (likely) benign variants for *BRCA1* and 92.2% and 86.6% for *BRCA2*. Notably for the majority of presumed LOF variants assigned evidence against pathogenicity are not assigned full or any pathogenic very strong (PVS1) code strength according to the recent ENIGMA classification criteria, since their predicted or known impact on splicing indicates that they are not associated with high risk of cancer^2,20^.

Using reference sets of (likely) pathogenic and (likely) benign variants drawn from ClinVar; we used the CDS±2bp since this captures the regions more regularly tested in a clinical setting, we showed that the ccLR method maintains low false discovery and false omission rates (< 0.05) and can reach strong evidence both in favor and against pathogenicity, following the Bayesian adaptation of the ACMG/AMP framework^4,5^. Our results provide case-control LR evidence for 1,453 variants with suggestive ACMG/AMP code strength levels; of these 266 have evidence in favor of pathogenicity, and 1,187 have evidence against pathogenicity. This analysis provides evidence to inform clinical classification for 785 variants currently considered of uncertain clinical significance.

While our study provides valuable results that can be used in variant classification assuming the variant follows a high-risk penetrance as observed for “average” *BRCA1* and *BRCA2* pathogenic variants. However, it is important to acknowledge possible sources of bias such as potential sequencing artifacts, variant allele frequency differences between populations, case-control imbalances and possibility of ascertainment bias. We addressed this by applying stringent quality control measures and stratified analyses. It is important to also acknowledge the fact that when assessing numerous variants, particularly those with low counts, some may exhibit outcomes contrary to the expected, by chance. This issue is particularly relevant in the Supporting evidence category, where evidence is relatively weak. However, by future integration of case-control LR evidence with other lines of evidence the likelihood of misclassification is minimized.

We further demonstrated that ccLR evidence aligns with functional predictors, offering a robust evaluation framework. These results can serve as a crucial resource for evaluating the consistency of other findings and refining existing methodologies.

In summary, we present case-control evidence that strongly aligns with ClinVar pathogenicity assertions for non-VUS. We also provide case-control evidence towards or against pathogenicity for 785 unclassified variants. This can now be used in combination with other evidence for their clinical classification, which is essential for accurate risk assessment and effective clinical decision-making, providing a larger number of patients and their relatives with clinically informative results.

## Methods

### The BCAC/BRIDGES dataset

The BCAC dataset included 47,201 women with breast cancer and 47,316 unaffected controls, from 29 BCAC studies defined as population-based^8^. All studies were approved by the respective ethics review boards, adhering to appropriate consent procedures. Phenotype data were based on the BCAC database v14. Individuals from a minority ancestry for each study (i.e., non-Asian from the Asian studies and non-European ancestry from the other studies), based on genetic data or self-report, were excluded^8^. Samples underwent panel sequencing for 34 known or suspected breast cancer susceptibility genes as part of the BRIDGES project^8^. Details on library preparation, sequencing and bioinformatics analysis including variant calling and quality control are described elsewhere^8^. The final dataset used consisted of 46,306 women with breast cancer and 43,481 unaffected controls, diagnosed or interviewed at age 21 to 80 years (the age range for which penetrance estimates for *BRCA1* and *BRCA2* are available) (**Fig. 8** and **Supplementary Table 5**).

**Fig. 8:**
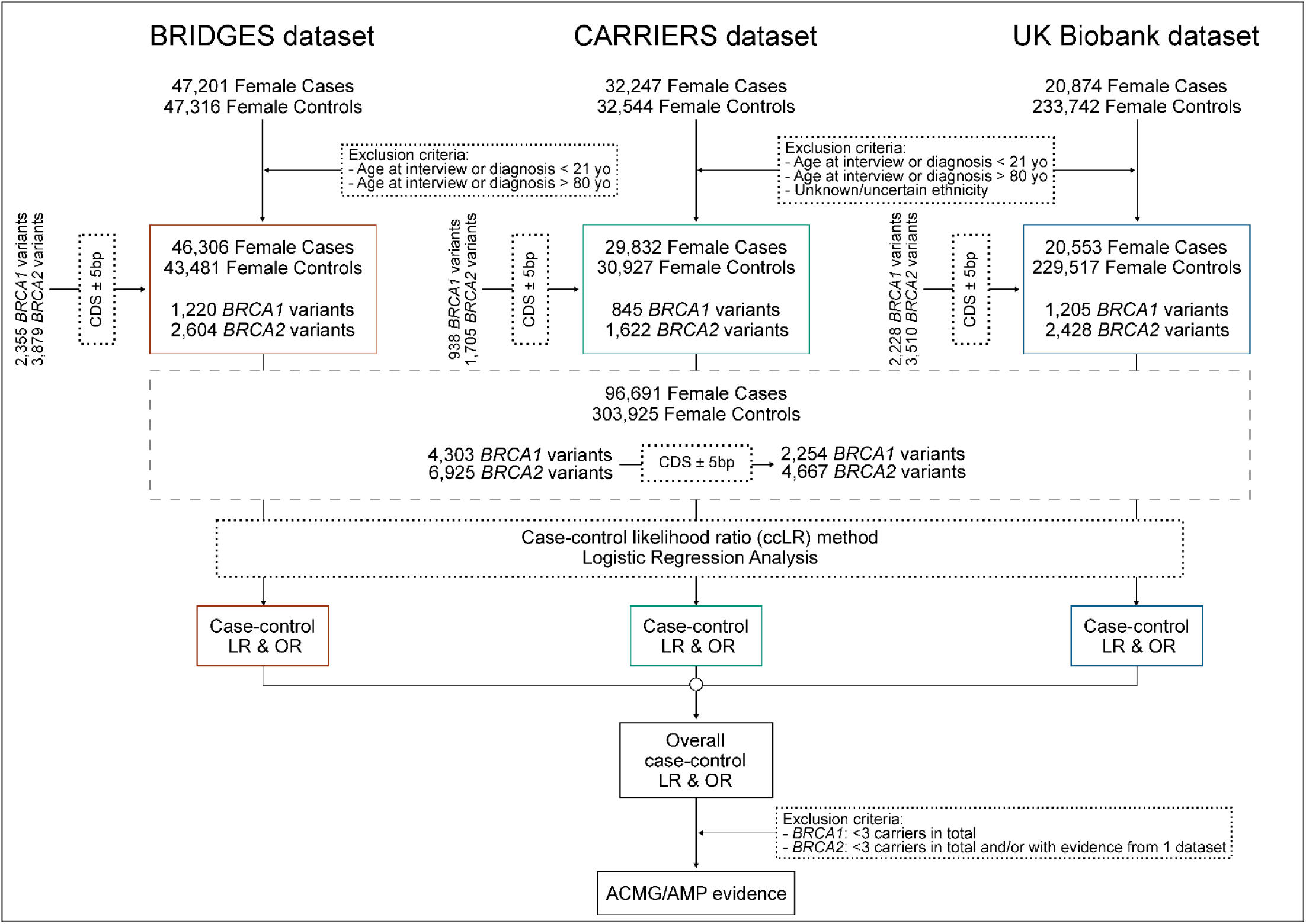
Flowchart summarizing the study design. Using sequencing data of 96,691 female breast cancer cases and 304,649 unaffected controls from the Breast Cancer Association Consortium (BCAC), the Cancer Risk Estimates Related to Susceptibility (CARRIERS) consortium and the UK Biobank (UKB) we calculated case-control likelihood ratios (LRs) and odds ratios (ORs) for 11,264 *BRCA1* and *BRCA2* variants, of which 6,943 are coding (coding sequence, CDS±5bp). Derived LRs and ORs were further aligned to ACMG/AMP evidence strengths to provide evidence in favor or against pathogenicity following sensitivity analyses-derived variant exclusion criteria

### The CARRIERS dataset

The CARRIERS consortium dataset included 32,247 women with breast cancer and 32,544 unaffected controls from 12 population-based studies^9^. The CARRIERS study was approved by the institutional review board at the Mayo Clinic and all participants provided informed consent for research. Samples were subjected to panel sequencing, targeting 37 cancer susceptibility genes. Details on library preparation, sequencing and bioinformatics analysis including variant calling and quality control were previously documented^9^. Women of unknown/uncertain ethnicity were excluded. The final dataset consisted of 29,832 women with breast cancer and 30,927 unaffected controls diagnosed or interviewed, between the age range of 21 to 80 years (**Fig. 8** and **Supplementary Table 5**).

### The UK Biobank dataset

UKB is a prospective cohort of more than 500,000 participants recruited in 22 assessment centers in the United Kingdom between 2006 and 2010^21^. Whole-exome sequencing (WES) data for 454,787 samples were released in October 2021 and were accessed via the UKB DNA Nexus platform^10^. Genetic ancestry was computed using a genetic principal components analysis from 2,318 informative markers^22^. Women of unknown/uncertain ethnicity were excluded. Cases were defined as individuals with either invasive breast cancer (International Classification of Diseases (ICD)-10 code (C50) or carcinoma in situ (D05) based on linkage to the National Cancer Registration and Analysis Service (CRAS), or self-reported breast cancer incidence. We included both prevalent and incident cases, and only breast cancers identified as an individual’s first or second diagnosed cancer. Under these criteria, a total of 20,553 female breast cancer cases and 229,517 unaffected female individuals were included, diagnosed or interviewed between the age range of 21 to 80 years (**Fig. 8** and **Supplementary Table 5**). Access to the use of the UK Biobank data was granted under application number 102655.

### Data preparation

For all datasets, the Ensembl Variant Effect Predictor (VEP) v111 was used to annotate variants^23^. Annotations include the distance from the upstream or downstream gene, sequence ontology variant consequences, exon/intron number and Human Genome Variation Society (HGVS) Nomenclature for the cDNA and protein level; the MANE Select transcript and protein were used (ENST00000357654.3 and ENSP00000350283.3 for *BRCA1*; ENST00000380152.3 and ENSP00000369497.3 for *BRCA2*)^24^. Exons and introns are sequentially numbered to match the MANE Select transcripts. Although *BRCA1* was initially described with 24 exons (GenBank Assession ID U14680.1), exon 4 is missing following further assessment of the gene. Herein, we implement the most updated version of exon numbering (excluding legacy exon numbering). Allele frequency (AF) was retrieved from the gnomAD v4.1.0 release^25^. For filtering variants, we used the gnomAD maximum credible AF (the lower bound of the 95% CI) observed across the non-founder populations, including non-Finnish Europeans, African or African Americans, Admixed Americans, East Asians, South Asians and Middle Easterners. Existing variant class was retrieved from the ClinVar database (https://www.ncbi.nlm.nih.gov/clinvar/, last accessed on January 7, 2024).

### Case-control analysis

The primary analysis involved the calculation of LRs for each individual variant using the ccLR method (https://github.com/BiostatUnitCING/ccLR)^7^. Under a survival analysis framework, the ccLR method compares the likelihood of the distribution of the variant of interest between cases and controls, under the hypothesis that the variant has similar age-specific relative risks as the “average” *BRCA1* or *BRCA2* pathogenic variant (PV), compared to the hypothesis that the variant is not associated with increased risk of the disease^7^. These risks are age-, sex-, and/or country-specific. Hence, the ccLR method requires specification of the age-specific risks in individuals with a PV and in the general population^7^. These were derived from the age-specific incidence rates for England and Wales (1998-2002) for ages 21-80 years, retrieved by the Cancer Incidence in Five Continents (CI5) Volume IX (https://ci5.iarc.fr/CI5I-X/Default.aspx) and age-specific breast cancer odds ratio for individuals with *BRCA1* and *BRCA2* PVs estimated from the BRIDGES study^8^.

Case-control LRs were separately calculated for each dataset: BRIDGES, CARRIERS and UKB. To account for possible variant allele frequency differences by country or ethnicity, stratified LR calculations were performed within each dataset (BRIDGES stratified by country, and hence largely also by ethnicity; CARRIERS and UKB stratified by ethnicity) (**Supplementary Table 6**) and then multiplied across strata to provide a case-control LR for each independent dataset. Dataset-specific LRs were then used to obtain an overall LR for each variant. The LRs were aligned to evidence strength levels for or against pathogenicity based on the thresholds recommended under the ACMG/AMG framework^4^. Thus, in favor of pathogenicity was classified as very strong, LR ≥ 350; strong, 350 > LR ≥ 18.7; moderate, 18.7 > LR ≥ 4.33; or supporting, 4.33 > LR ≥ 2.08. Likelihood ratios against pathogenicity were classified as very strong, LR ≤ 0.0029; strong, 0.0029 < LR ≤ 0.053; moderate, 0.053 < LR ≤ 0.231; and supporting, 0.231 < LR ≤ 0.48. LRs between 0.48 and 2.08 were considered of “No evidence”.

Associations between variants and breast cancer risk were also assessed by logistic regression, adjusted for age and study country for the BCAC dataset, age and ethnic group for the CARRIERS dataset, and age and genetic ancestry for the UKB dataset. Logistic regression analysis was only performed for variants present in both cases and controls. Odds ratios and standard errors estimated from each dataset were combined in a fixed-effects, inverse-variance meta-analysis using the ‘*metafor’ R* package to derive an overall test of association. Using the *BRCA1/2* VCEP specification based on the PS4 ACMG/AMP classification criterion^2^, strong evidence in favor of pathogenicity was assigned to variants with OR ≥ 4.0, *P* value < 0.05 and CI not including 2.0.

### Reference Sets

To assess the calibration of the ccLR method and perform sensitivity analyses, we selected reference sets of (likely) pathogenic and (likely) benign variants from the list of *BRCA1* and *BRCA2* variants identified in any of the three datasets. These reference sets included variants located within CDS or at splice acceptor/donor site dinucleotide positions (±2bp); since this captures the regions more regularly tested in a clinical setting, classified as such by the ClinGen *BRCA1/2* historical expert panel or the ClinGen *BRCA1/2* VCEP following ACMG/AMP guidelines^2^, or by multiple submitters without conflicts in the ClinVar database (last accessed on January 7, 2024). Variants were filtered to exclude any with gnomAD (v4.1.0) maximum credible AF in non-founder populations, denoted as FAF > 0.001, consistent with the *BRCA1* and *BRCA2* VCEP’s “BA1” benign stand-alone cutoff. Reference sets included 739 *BRCA1* variants (448, 369 and 373 with case-control LRs from BRIDGES, CARRIERS and UKB, respectively) and 1,241 *BRCA2* variants (762, 554 and 634 with case-control LRs from BRIDGES, CARRIERS and UKB, respectively).

### Calibration of the ccLR method

To evaluate the calibration of the method, we used the case-control LRs for each of the variants in the reference sets and then determined the proportion of benign and pathogenic variants that would be classified in each ACMG/AMP evidence strength category^4^. We calculated enrichment for variants in each category *j* to be pathogenic as the ratio of proportions of variants in category *j* that are pathogenic or benign relative to the overall ratio of pathogenic and benign proportions^26^, that is:

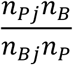

where *n_Pj_* and *n_Bj_* are the number of pathogenic and benign variants in category *j*, and *n_p_* and *n_B_* are the total number of pathogenic and benign variants.

For calibration purposes, instances where variants were assigned either very strong or strong evidence according to the ACMG/AMP criteria, either in favor or against pathogenicity were collectively considered as instances of strong evidence in favor of or against pathogenicity, respectively. The Haldane-Anscombe correction was applied for any category where the cell count for pathogenic or benign variant reference set was zero^27^.

### Concordance with predicted or experimental impact on function

To assess the concordance of the ccLR method with high-throughput functional assays and *in silico* prediction methods, we compared case-control LR values and suggestive ACMG/AMP code strength levels for variants located within CDS or at splice acceptor/donor site dinucleotide positions (±2bp); since this captures the regions more regularly tested in a clinical setting, present in at least three individuals in the combined dataset, and with evidence from at least two datasets for *BRCA2*.

Case-control evidence was compared with high-throughput functional assays performed for functionally critical *BRCA1* and *BRCA2* domains^11–13^ and *in silico* prediction methods. *In silico* prediction methods included BayesDel^14^ (without minor allele frequency) which is currently recommended (with optimal score cutpoints) by the ClinGen BRCA1/2 VCEP for the curation of *BRCA1* and *BRCA2* variants (benign BP4 criterion “Multiple lines of computational evidence suggest no impact on gene or gene product”, ≤ 0.15 for *BRCA1* and ≤ 0.18 for *BRCA2*; pathogenic PP3 criterion “Multiple lines of computational evidence support a deleterious effect on the gene or gene product”, ≥ 0.28 for *BRCA1* and ≥ 0.30 for *BRCA2*)^2^, as well as other computational tools recommended (with appropriate score thresholds) by the ClinGen SVI Working Group^28^, including REVEL^15^ (BP4, ≤ 0.29; PP3, ≥ 0.644), VEST4^16^ (BP4, ≤ 0.449; PP3, ≥ 0.764) and MutPred2^17^ (BP4, ≤ 0.391; PP3, ≥ 0.737). We also investigated the recent proteome-wide variant effect predictor AlphaMissense^18^.

Sensitivity and specificity metrics were used to assess the concordance of the functional predictors at predicting variants assigned evidence in favor or against pathogenicity using the ccLR method.

## Data Availability

Individual level data for the BCAC data are not publicly available due to ethical review board constraints but are available on request through the BCAC Data Access Co-ordinating Committee. Requests for access to UK Biobank data should be made to the UK Biobank Access Management Team. According to the "UK Biobank best practice approaches on the publication of small numbers, Tables and/or results of any UK Biobank data there should have a minimum number of 5 reported participants within a cell". Therefore, for variants present in less than 5 individuals, carrier count (Carriers, N) was replaced by "<5".

## Web resources

ClinVar database, https://www.ncbi.nlm.nih.gov/clinvar/

gnomAD database, https://gnomad.broadinstitute.org/

ccLR GitHub repository, https://github.com/BiostatUnitCING/ccLR

Cancer Incidence in Five Continents (CI5) Volume IX, https://ci5.iarc.fr/CI5I-X/Default.aspx

LiftOver tool, https://genome.ucsc.edu/cgi-bin/hgLiftOver

## Acknowledgments

This research has been conducted using the UK Biobank Resource under application number 102655. Access to the UK Biobank data has been funded by an internal research grant of the Cyprus Institute of Neurology and Genetics. The BRIDGES panel sequencing was supported by the European Union Horizon 2020 research and innovation program BRIDGES (grant number, 634935) and the Wellcome Trust (v203477/Z/16/Z). The CARRIERS studies were supported in part by NIH grants R35CA253187, R01CA225662, P50CA116201 and the Breast Cancer Research Foundation (BCRF). Details on individual and study funding and acknowledgments are in **Supplementary File 1**.

## Author Contributions

M.Z., D.G.O., M.T.P., K.M., A.B.S. and D.F.E., drafted the manuscript; K.M., A.B.S., D.F.E., F.J.C, D.E.G., M.Z. and D.G.O. designed the study. M.Z., D.O.M., K.Y., K.M., conducted statistical analyses. M.Z., L.D., J.D., M.K.B., Q.W., N.J.B., W.C., C.H., M.N. conducted bioinformatics analyses and data collection. T.U.A., C.B.A., I.L. A., A.C.A., P.L.A., C.B., C.B., N.V.B., S.E.B., M.K.B., K.D.B., N.J.C., A.C., J.E.C., M.H.C., J.C-C., F.C., G.C-T., NBCS Collaborators, D.M.C., K.C., S.M.D., T.D., A.M.D., A.H.E., D.G.E., P.A.F., J.D.F., H.F., M.G-D., M.G-C., G.G., A.G-N., F.G., A.H., C.A.H., U.H., S.N.H., M.B.A.H, W-K.H., J.M.H., R.H., S.J.H., C.H., kConFab Investigators, A.J., E.K.K., Y-D.K., P.K., V.N.K., J.V.L., J.L., G.H.L., S.L., A.L., C.L., A.M., M.E.M., D.M., R.L.M., K.M., K.L.N., R.N-T., N.O., J.E.O., J.R.P., M.I.P., A.V.P., P.D.P.P., E.C.P., M.U.R., K.J.R., E.S., E.J.S., M.K.S., M.C.S., V.K-M.T., S.H.T., L.R.T., D.T., A.T-D., T.T., C.M.V., Q.W., J.N.W., S.Y., S.Y., G.R.Z. provided data. All authors contributed to review and editing. All authors read and approved the final version of the manuscript.

