## Supplementary File 1 for "Analysis of more than 400,000 women provides case-control evidence for BRCA1 and BRCA2 variant classification"

**Supplementary File 1.** Individual and study funding and acknowledgments

**Funding**

This work was supported by Cancer Research UK grant: PPRPGM-Nov20\100002 and by core funding from the NIHR Cambridge Biomedical Research Centre (NIHR203312) [*]. *The views expressed are those of the author(s) and not necessarily those of the NIHR or the Department of Health and Social Care. Additional funding for BCAC is provided by the Confluence project which is funded with intramural funds from the National Cancer Institute Intramural Research Program, National Institutes of Health, the European Union's Horizon 2020 Research and Innovation Programme (grant numbers 634935 and 633784 for BRIDGES and B-CAST respectively), and the PERSPECTIVE I&I project, funded by the Government of Canada through Genome Canada and the Canadian Institutes of Health Research, the Ministère de l’Économie et de l'Innovation du Québec through Genome Québec, the Quebec Breast Cancer Foundation. The EU Horizon 2020 Research and Innovation Programme funding source had no role in study design, data collection, data analysis, data interpretation or writing of the report.

ABS was supported by an NHMRC Investigator Fellowship (APP177524). The work of MTP was supported in part by National Institutes of Health grant U24 5U24CA258058-02. SVT’s analytical contribution was funded by NCI grant R01CA264971 (“Upgrading rigor and efficiency of germline cancer gene variant classification for the 2020s”). PK's contribution was supported by the Intramural Research Program of the National Cancer Institute. MSC was supported by the National Institute of Health Grants R01CA264971 and U24CA258058.

The ABCS study was supported by the Dutch Cancer Society [grants NKI 2007-3839; 2009 4363] and an institutional grant of the Dutch Cancer Society and of the Dutch Ministry of Health, Welfare and Sport. The ACP study is funded by the Breast Cancer Research Trust, UK. KM and AL are supported by the NIHR Manchester Biomedical Research Centre, the Allan Turing Institute under the EPSRC grant EP/N510129/1. The work of the BBCC was partly funded by ELAN-Fond of the University Hospital of Erlangen. For BIGGS, ES is supported by NIHR Comprehensive Biomedical Research Centre, Guy's & St. Thomas' NHS Foundation Trust in partnership with King's College London, United Kingdom. IT is supported by the Oxford Biomedical Research Centre. BOCS is supported by funds from Cancer Research UK (C8620/A8372/A15106) and the Institute of Cancer Research (UK). The BREast Oncology GAlician Network (BREOGAN) is funded by Acción Estratégica de Salud del Instituto de Salud Carlos III FIS PI12/02125/Cofinanciado and FEDER PI17/00918/Cofinanciado FEDER; Acción Estratégica de Salud del Instituto de Salud Carlos III FIS Intrasalud (PI13/01136); Programa Grupos Emergentes, Cancer Genetics Unit, Instituto de Investigacion Biomedica Galicia Sur. Xerencia de Xestion Integrada de Vigo-SERGAS, Instituto de Salud Carlos III, Spain; Grant 10CSA012E, Consellería de Industria Programa Sectorial de Investigación Aplicada, PEME I + D e I + D Suma del Plan Gallego de Investigación, Desarrollo e Innovación Tecnológica de la Consellería de Industria de la Xunta de Galicia, Spain; Grant EC11-192. Fomento de la Investigación Clínica Independiente, Ministerio de Sanidad, Servicios Sociales e Igualdad, Spain; and Grant FEDER-Innterconecta. Ministerio de Economia y Competitividad, Xunta de Galicia, Spain. The BSUCH study was supported by the Dietmar-Hopp Foundation, the Helmholtz Society and the German Cancer Research Center (DKFZ). CCGP is supported by funding from the University of Crete. The CECILE study was supported by Fondation de France, Institut National du Cancer (INCa), Ligue Nationale contre le Cancer, Agence Nationale de Sécurité Sanitaire, de l'Alimentation, de l'Environnement et du Travail (ANSES), Agence Nationale de la Recherche (ANR). The CGPS was supported by the Chief Physician Johan Boserup and Lise Boserup Fund, the Danish Medical Research Council, and Herlev and Gentofte Hospital. The CNIO-BCS was supported by the Instituto de Salud Carlos III, the Red Temática de Investigación Cooperativa en Cáncer and grants from the Asociación Española Contra el Cáncer and the Fondo de Investigación Sanitario (PI11/00923 and PI12/00070). COLBCCC is supported by the German Cancer Research Center (DKFZ), Heidelberg, Germany. Diana Torres was in part supported by a postdoctoral fellowship from the Alexander von Humboldt Foundation. PROCAS is funded from NIHR grant PGfAR 0707-10031. DGE, AH and WGN are supported by the NIHR Manchester Biomedical Research Centre (IS-BRC-1215-20007). The GENICA was funded by the Federal Ministry of Education and Research (BMBF) Germany grants 01KW9975/5, 01KW9976/8, 01KW9977/0 and 01KW0114, the Robert Bosch Foundation, Stuttgart, Deutsches Krebsforschungszentrum (DKFZ), Heidelberg, the Institute for Prevention and Occupational Medicine of the German Social Accident Insurance, Institute of the Ruhr University Bochum (IPA), Bochum, as well as the Department of Internal Medicine, Johanniter GmbH Bonn, Johanniter Krankenhaus, Bonn, Germany. Generation Scotland (GENSCOT) received core support from the Chief Scientist Office of the Scottish Government Health Directorates [CZD/16/6] and the Scottish Funding Council [HR03006]. The GESBC was supported by the Deutsche Krebshilfe e. V. [70492] and the German Cancer Research Center (DKFZ). The HABCS study was supported by German Research Foundation (DFG Do761/15-1), the Claudia von Schilling Foundation for Breast Cancer Research, by the Lower Saxonian Cancer Society, and by the Rudolf Bartling Foundation. The HMBCS was supported by the German Research Foundation (DFG Do761/15-1), a grant from the Friends of Hannover Medical School, and by the Rudolf Bartling Foundation. The HUBCS was supported by German Research Foundation (DFG Do761/15-1), a grant from the German Federal Ministry of Research and Education (RUS08/017), B.M. was supported by grant 17-44-020498, 17-29-06014 of the Russian Foundation for Basic Research, D.P. was supported by grant 18-29-09129 of the Russian Foundation for Basic Research, E.K was supported by the mega grant from the Government of Russian Federation (2020-220-08-2197), and the study was performed as part of the assignment of the Ministry of Science and Higher Education of the Russian Federation (№АААА-А16-116020350032-1). Financial support for KARBAC was provided through the regional agreement on medical training and clinical research (ALF) between Stockholm County Council and Karolinska Institutet, the Swedish Cancer Society, The Gustav V Jubilee foundation and Bert von Kantzows foundation. The KARMA study was supported by Märit and Hans Rausings Initiative Against Breast Cancer. The KBCP was financially supported by the special Government Funding (VTR) of Kuopio University Hospital grants, Cancer Fund of North Savo, the Finnish Cancer Organizations, and by the strategic funding of the University of Eastern Finland. kConFab is supported by a grant from the National Breast Cancer Foundation, and previously by the National Health and Medical Research Council (NHMRC), the Queensland Cancer Fund, the Cancer Councils of New South Wales, Victoria, Tasmania and South Australia, and the Cancer Foundation of Western Australia. Financial support for the AOCS was provided by the United States Army Medical Research and Materiel Command [DAMD17-01-1-0729], Cancer Council Victoria, Queensland Cancer Fund, Cancer Council New South Wales, Cancer Council South Australia, The Cancer Foundation of Western Australia, Cancer Council Tasmania and the National Health and Medical Research Council of Australia (NHMRC; 400413, 400281, 199600). G.C.T. and P.W. are supported by the NHMRC. RB was a Cancer Institute NSW Clinical Research Fellow. The MARIE study was supported by the Deutsche Krebshilfe e.V. [70-2892-BR I, 106332, 108253, 108419, 110826, 110828], the Hamburg Cancer Society, the German Cancer Research Center (DKFZ) and the Federal Ministry of Education and Research (BMBF) Germany [01KH0402]. The MASTOS study was supported by “Cyprus Research Promotion Foundation” grants 0104/13 and 0104/17, and the Cyprus Institute of Neurology and Genetics. The Melbourne Collaborative Cohort Study (MCCS) cohort recruitment was funded by VicHealth and Cancer Council Victoria. The MCCS was further augmented by Australian National Health and Medical Research Council grants 209057, 396414 and 1074383 and by infrastructure provided by Cancer Council Victoria. Cases and their vital status were ascertained through the Victorian Cancer Registry and the Australian Institute of Health and Welfare, including the National Death Index and the Australian Cancer Database. MYBRCA is funded by research grants from the Wellcome Trust (v203477/Z/16/Z), the Malaysian Ministry of Higher Education (UM.C/HlR/MOHE/06) and Cancer Research Malaysia. The NBCS has received funding from the K.G. Jebsen Centre for Breast Cancer Research; the Research Council of Norway grant 193387/V50 (to A-L Børresen-Dale and V.N. Kristensen) and grant 193387/H10 (to A-L Børresen-Dale and V.N. Kristensen), South Eastern Norway Health Authority (grant 39346 to A-L Børresen-Dale) and the Norwegian Cancer Society (to A-L Børresen-Dale and V.N. Kristensen). The Ontario Familial Breast Cancer Registry (OFBCR) were supported by grant U01CA164920 from the USA National Cancer Institute of the National Institutes of Health. The PBCS was funded by Intramural Research Funds of the National Cancer Institute, Department of Health and Human Services, USA. Genotyping for PLCO was supported by the Intramural Research Program of the National Institutes of Health, NCI, Division of Cancer Epidemiology and Genetics. The PLCO is supported by the Intramural Research Program of the Division of Cancer Epidemiology and Genetics and supported by contracts from the Division of Cancer Prevention, National Cancer Institute, National Institutes of Health. The POSH study is funded by Cancer Research UK (grants C1275/A11699, C1275/C22524, C1275/A19187, C1275/A15956 and Breast Cancer Campaign 2010PR62, 2013PR044. The SASBAC study was supported by funding from the Agency for Science, Technology and Research of Singapore (A*STAR), the US National Institute of Health (NIH) and the Susan G. Komen Breast Cancer Foundation. The SBCGS was supported primarily by NIH grants R01CA64277, R01CA148667, UMCA182910, and R37CA70867. Biological sample preparation was conducted the Survey and Biospecimen Shared Resource, which is supported by P30 CA68485. The scientific development and funding of this project were, in part, supported by the Genetic Associations and Mechanisms in Oncology (GAME-ON) Network U19 CA148065. SEARCH was funded by Cancer Research UK [C490/A10124, C490/A16561] and supported by the UK National Institute for Health Research Biomedical Research Centre at the University of Cambridge. SGBCC is funded by the National Research Foundation Singapore, NUS start-up Grant, National University Cancer Institute Singapore (NCIS) Centre Grant, Breast Cancer Prevention Programme, Asian Breast Cancer Research Fund, Breast Cancer Screening and Prevention Programme and the NMRC Clinician Scientist Award (SI Category). Population-based controls were from the Multi-Ethnic Cohort (MEC) funded by grants from the Ministry of Health, Singapore, National University of Singapore and National University Health System, Singapore. SKKDKFZS is supported by the DKFZ. The SZBCS was supported by Grant PBZ_KBN_122/P05/2004 and the program of the Minister of Science and Higher Education under the name "Regional Initiative of Excellence" in 2019-2022 project number 002/RID/2018/19 amount of financing 12 000 000 PLN. UBCS was supported by funding from National Cancer Institute (NCI) grant R01 CA163353 (to N.J. Camp) and the Women’s Cancer Center at the Huntsman Cancer Institute (HCI). Data collection for UBCS was supported by the Utah Population Database (UPDB) and Utah Cancer Registry (UCR). The UPDB is supported by HCI (including Huntsman Cancer Foundation, HCF), the University of Utah, and NCI grant P30 CA2014. The UCR is funded by the NCI's SEER Program, Contract No. HHSN261201800016I, the US Center for Disease Control and Prevention's National Program of Cancer Registries (Cooperative Agreement No. NU58DP007131), the University of Utah, and HCF. The BWHS study was supported by NIH grants U01CA164974 and R01CA098663, as well as the Susan G. Komen Foundation grant SAC220228. The NHS was supported by NIH grants P01 CA87969, UM1 CA186107, and U19 CA148065. The NHS2 was supported by NIH grants UM1 CA176726 and U19 CA148065. The CTS and the research reported in this publication were supported by the National Cancer Institute of the National Institutes of Health under award number U01-CA199277; P30-CA033572; P30-CA023100; UM1-CA164917; and R01-CA077398. The collection of cancer incidence data used in the California Teachers Study was supported by the California Department of Public Health pursuant to California Health and Safety Code Section 103885; Centers for Disease Control and Prevention's National Program of Cancer Registries, under cooperative agreement 5NU58DP006344; the National Cancer Institute's Surveillance, Epidemiology and End Results Program under contract HHSN2612018000321 awarded to the University of California, San Francisco, contract HHSN2612018000151 awarded to the University of Southern California, and contract HHSN2612018000091 awarded to the Public Health Institute. The opinions, findings, and conclusions expressed herein are those of the author(s) and do not necessarily reflect the official views of the State of California, Department of Public Health, the National Cancer Institute, the National Institutes of Health, the Centers for Disease Control and Prevention or their Contractors and Subcontractors, or the Regents of the University of California, or any of its programs. The MEC study was supported by the NIH grant U01-CA164973. The WCHS was supported by the NCI grants R01 CA100598 and R01 CA185623. Wisconsin Women’s Health Study (WWHS) is supported by NCI grant P30CA014520 to the University of Wisconsin Carbone Cancer Center. The American Cancer Society funds the creation, maintenance, and updating of the Cancer Prevention Study-II Cancer Prevention Study-3 cohorts. CB, JH, AP, and LT's contribution was supported by American Cancer Society intramural research funding.

**Acknowledgements**

We thank all the individuals who took part in these studies and all the researchers, clinicians, technicians and administrative staff who have enabled this work to be carried out.

ABCS thanks the Blood bank Sanquin, The Netherlands. ABCTB Investigators: Christine Clarke, Deborah Marsh, Rodney Scott, Robert Baxter, Desmond Yip, Jane Carpenter, Alison Davis, Nirmala Pathmanathan, Peter Simpson, J. Dinny Graham, Mythily Sachchithananthan. Samples are made available to researchers on a non-exclusive basis. The ACP study wishes to thank the participants in the Thai Breast Cancer study. Special thanks also go to the Thai Ministry of Public Health (MOPH), doctors and nurses who helped with the data collection process. Finally, the study would like to thank Dr Prat Boonyawongviroj, the former Permanent Secretary of MOPH and Dr Pornthep Siriwanarungsan, the former Department Director-General of Disease Control who have supported the study throughout. BIGGS thanks Niall McInerney, Gabrielle Colleran, Andrew Rowan, Angela Jones. The BREOGAN study would not have been possible without the contributions of the following: Manuela Gago-Dominguez, Jose Esteban Castelao, Angel Carracedo, Victor Muñoz Garzón, Alejandro Novo Domínguez, Maria Elena Martinez, Sara Miranda Ponte, Carmen Redondo Marey, Maite Peña Fernández, Manuel Enguix Castelo, Maria Torres, Manuel Calaza (BREOGAN), José Antúnez, Máximo Fraga and the staff of the Department of Pathology and Biobank of the University Hospital Complex of Santiago-CHUS, Instituto de Investigación Sanitaria de Santiago, IDIS, Xerencia de Xestion Integrada de Santiago-SERGAS; Joaquín González-Carreró and the staff of the Department of Pathology and Biobank of University Hospital Complex of Vigo, Instituto de Investigacion Biomedica Galicia Sur, SERGAS, Vigo, Spain. The BSUCH study acknowledges the Principal Investigator, Barbara Burwinkel, and, thanks Peter Bugert, Medical Faculty Mannheim. CCGP thanks Styliani Apostolaki, Anna Margiolaki, Georgios Nintos, Maria Perraki, Georgia Saloustrou, Georgia Sevastaki, Konstantinos Pompodakis. CGPS thanks staff and participants of the Copenhagen General Population Study. For the excellent technical assistance: Dorthe Uldall Andersen, Maria Birna Arnadottir, Anne Bank, Dorthe Kjeldgård Hansen. CNIO-BCS thanks Guillermo Pita, Charo Alonso, Nuria Álvarez, Pilar Zamora, Primitiva Menendez, the Human Genotyping-CEGEN Unit (CNIO). COLBCCC thanks all patients, the physicians Justo G. Olaya, Mauricio Tawil, Lilian Torregrosa, Elias Quintero, Sebastian Quintero, Claudia Ramírez, José J. Caicedo, and Jose F. Robledo, and the technician Michael Gilbert for their contributions and commitment to this study. PROCAS thanks NIHR for funding. The GENICA Network: Dr. Margarete Fischer-Bosch-Institute of Clinical Pharmacology, Stuttgart, and University of Tübingen, Germany [Hiltrud Brauch, RH, Wing-Yee Lo], Department of Internal Medicine, Johanniter GmbH Bonn, Johanniter Krankenhaus, Bonn, Germany [YDK, Christian Baisch], Institute of Pathology, University of Bonn, Germany [Hans-Peter Fischer], Molecular Genetics of Breast Cancer, Deutsches Krebsforschungszentrum (DKFZ), Heidelberg, Germany [UH], Institute for Prevention and Occupational Medicine of the German Social Accident Insurance, Institute of the Ruhr University Bochum (IPA), Bochum, Germany [Thomas Brüning, Beate Pesch, Sylvia Rabstein, Anne Lotz]; and Institute of Occupational Medicine and Maritime Medicine, University Medical Center Hamburg-Eppendorf, Germany [Volker Harth]. HABCS thanks Peter Schürmann, Peter Hillemanns, Natalia Bogdanova, Michael Bremer, Johann Karstens, Hans Christiansen and the Breast Cancer Network in Lower Saxony for continuous support. HMBCS thanks Peter Hillemanns, Hans Christiansen and Johann H. Karstens. HUBCS thanks Darya Prokofyeva and Shamil Gantsev. ICICLE thanks Kelly Kohut, Michele Caneppele, Maria Troy. KARMA and SASBAC thank the Swedish Medical Research Counsel. KBCP thanks Eija Myöhänen. kConFab/AOCS wish to thank Heather Thorne, Eveline Niedermayr, all the kConFab research nurses and staff, the heads and staff of the Family Cancer Clinics, and the Clinical Follow Up Study (which has received funding from the NHMRC, the National Breast Cancer Foundation, Cancer Australia, and the National Institute of Health (USA)) for their contributions to this resource, and the many families who contribute to kConFab. MARIE thanks Petra Seibold, Nadia Obi, Sabine Behrens, Ursula Eilber and Muhabbet Celik. MASTOS thanks all the study participants and express appreciation to the doctors: Yiola Marcou, Eleni Kakouri, Panayiotis Papadopoulos, Simon Malas and Maria Daniel, as well as to all the nurses and volunteers who provided valuable help towards the recruitment of the study participants. The MCCS was made possible by the contribution of many people, including the original investigators, the teams that recruited the participants and continue working on follow-up, and the many thousands of Melbourne residents who continue to participate in the study. MYBRCA thanks study participants and research staff (particularly Patsy Ng, Nurhidayu Hassan, Yoon Sook-Yee, Daphne Lee, Lee Sheau Yee, Phuah Sze Yee and Norhashimah Hassan) for their contributions and commitment to this study. The following are NBCS Collaborators: Kristine K. Sahlberg (PhD), Anne-Lise Børresen-Dale (Prof. Em.), Lars Ottestad (MD), Rolf Kåresen (Prof. Em.) Dr. Ellen Schlichting (MD), Marit Muri Holmen (MD), Toril Sauer (MD), Vilde Haakensen (MD), Olav Engebråten (MD), Bjørn Naume (MD), Alexander Fosså (MD), Cecile E. Kiserud (MD), Kristin V. Reinertsen (MD), Åslaug Helland (MD), Margit Riis (MD), Jürgen Geisler (MD), OSBREAC and Grethe I. Grenaker Alnæs (MSc). NBHS and SBCGS thank study participants and research staff for their contributions and commitment to the studies. The OFBCR thanks Teresa Selander, Nayana Weerasooriya and Steve Gallinger. ORIGO thanks E. Krol-Warmerdam, and J. Blom for patient accrual, administering questionnaires, and managing clinical information. PBCS thanks Louise Brinton, Mark Sherman, Neonila Szeszenia-Dabrowska, Beata Peplonska, Witold Zatonski, Pei Chao, Michael Stagner. We thank the SEARCH team. SGBCC thanks the participants and all research coordinators for their excellent help with recruitment, data and sample collection. SKKDKFZS thanks all study participants, clinicians, family doctors, researchers and technicians for their contributions and commitment to this study. UBCS thanks all study participants as well as the ascertainment, laboratory, analytics and informatics teams at Huntsman Cancer Institute and Intermountain Healthcare for their important contributions to this study. BWHS thanks the contributions of central cancer registries supported through the Centers for Disease Control and Prevention National Program of Cancer Registries (NPCR) and/or the National Cancer Institute Surveillance, Epidemiology, and End Results (SEER) Program. Central registries may also be supported by state agencies, universities, and cancer centers. Participating central cancer registries include the following: AL, AR, AZ, CA, CO, CT, DE, DC, FL, GA, HI, IA, IL, IN, KY, LA, MD, MA, MI, MO, MS, NE, NJ, NM, NY, NC, OH, OK, OR, PA, SC, TN, TX, VA, WA, WI. The content is solely the responsibility of the authors and does not necessarily represent the official views of the U.S. Department of Health and Human Services, the National Institutes of Health, the National Cancer Institute, or the state cancer registries. WHI would like to also acknowledge the following short list of WHI investigators: Program Office: (National Heart, Lung, and Blood Institute, Bethesda, Maryland) Jacques Rossouw, Shari Ludlam, Dale Burwen, Joan McGowan, Leslie Ford, and Nancy Geller. WHI Clinical Coordinating Center (Fred Hutchinson Cancer Research Center, Seattle WA) and the WHI investigators (Garnet Anderson, Ross Prentice, Andrea LaCroix, Charles Kooperberg, JoAnn E. Manson, Barbara V. Howard, Marcia L. Stefanick, Rebecca Jackson, Cynthia A. Thomson, Jean Wactawski-Wende, Marian Limacher, Jennifer Robinson, Lewis Kuller, Sally Shumaker, Robert Brunner, Karen L. Margolis) for their dedication, and the study participants for making the program possible. A full list of WHI investigators can be found at: https://www.whi.org/researchers/Documents%20%20Write%20a%20Paper/WHI%20Investigator%20Long%20List.pdf. The Women’s Health Initiative (WHI) programme is funded by the National Heart, Lung, and Blood Institute, National Institutes of Health, US Department of Health and Human Services through contracts 75N92021D00001, 75N92021D00002, 75N92021D00003, 75N92021D00004, 75N92021D00005. The authors of the CTS would like to thank the CTS Steering Committee that is responsible for the formation and maintenance of the Study within which this research was conducted. A full list of California Teachers Study team members is available at <https://www.calteachersstudy.org/team>. The CPS3 and CPSII authors express sincere appreciation to all Cancer Prevention Study-II and Cancer Prevention Study-3 participants, and to each member of the study and biospecimen management group. The authors would also like to acknowledge the contribution to this study from central cancer registries supported through the Centers for Disease Control and Prevention's National Program of Cancer Registries and cancer registries supported by the National Cancer Institute's Surveillance Epidemiology and End Results Program. WWHS thanks Elizabeth Burnside, Irene Ong, John Hampton, and Julie McGregor for study support.
